# Association between Growth Differentiation Factor-15 and Coagulation Parameters in Male Chinese Coronary Artery Disease Patients

**DOI:** 10.1101/2025.05.08.25327206

**Authors:** Huan Liu, Yongnan Lyu, Wen Dai, Yan Li

**Affiliations:** Department of Clinical Laboratory, Institute of Translational Medicine, Renmin Hospital of Wuhan University, Wuhan, Hubei, PR China; Department of Cardiology, Renmin Hospital of Wuhan University, Wuhan, Hubei, PR China

**Keywords:** Growth differentiation factor-15, Coronary artery disease, Coagulation, Acute coronary syndrome

## Abstract

**Objectives:** Growth differentiation factor-15 (GDF-15) has emerged as a novel biomarker for coronary artery disease (CAD). Although a hypercoagulable state is recognized as a biological mechanism triggering cardiac events in CAD, the relationship between GDF-15 and coagulation parameters in CAD patients remains unclear.

**Methods:** We screened male patients who underwent elective coronary angiography for suspected CAD at Renmin Hospital of Wuhan University between January 2020 and December 2020, and enrolled those with confirmed diagnoses. Serum GDF-15 levels, blood cell counts, glucose, serum lipids, and coagulation parameters were measured.

**Results:** A total of 892 subjects were included (592 CAD patients and 300 controls). The CAD group comprised 161 stable angina (SA) and 431 acute coronary syndrome (ACS) cases. Data were analyzed using Kruskal–Wallis one-way ANOVA with post hoc tests (Holm–Sidak and Dunn’s tests). Compared with controls, CAD patients showed significantly higher serum GDF-15 levels. When stratified by GDF-15 tertiles, patients with high GDF-15 levels exhibited prolonged prothrombin time (PT) and activated partial thromboplastin time (APTT), elevated fibrinogen and D-dimer levels, and reduced antithrombin III (AT3) activity (all *P*<0.01). Multiple linear regression revealed that GDF-15 was independently associated with fibrinogen in all CAD patients (*β*=0.208, *P*<0.001) as well as in SA (*β*=0.171, *P*=0.035) and ACS subgroups (*β*=0.163, *P*=0.006). Additionally, GDF-15 negatively correlated with AT3 in the ACS subgroup (*β*= −0.120, *P*=0.039).

**Conclusions:** Elevated GDF-15 levels in male CAD patients are associated with altered coagulation parameters, suggesting that GDF-15 may serve as a potential indicator of thrombosis risk in cardiovascular management.

## Introduction

Coronary artery disease (CAD), the leading cause of death worldwide, is characterized by reversible myocardial ischemia due to demand/supply mismatch (1). Acute coronary syndromes (ACS), the primary clinical manifestation of coronary atherosclerosis, typically result from plaque rupture and subsequent thrombus formation in epicardial arteries, leading to acute occlusion (2). Thrombosis is central to ACS pathophysiology, driven by interrelated mechanisms such as endothelial dysfunction, inflammation, and coagulation. Systemic inflammation, for instance, promotes a proatherogenic state by upregulating prothrombotic factors and cell adhesion molecules, thereby activating platelets and facilitating clot formation (3). Thus, coagulation status critically influences CAD development and progression.

Growth differentiation factor-15 (GDF-15), a member of the transforming growth factor-β superfamily (4, 5), is markedly upregulated in pathological conditions like inflammation, cardiac injury, and oxidative stress (6–8). Emerging evidence highlight its role as a biomarker in cardiovascular diseases, including CAD (9–11), and suggests link to thrombosis. In early atherosclerosis, GDF-15 recruits macrophage to plaques (12), while its deficiency enhances plaque stability by reducing macrophage migration and increasing collagen deposition (13). Notably, GDF-15 independently predicts bleeding risk in atrial fibrillation (14–16) and correlates with venous thromboembolism risk and thrombus burden in deep venous thrombosis (17, 18). However, direct evidence linking GDF-15 to coagulation parameters in CAD remains scarce.

GDF-15’s prognostic value in CAD—predicting mortality and disease progression (9)—makes it clinically relevant (19, 20). Concurrently, assessing coagulation status is vital for thrombotic/bleeding risk management. Elevated GDF-15 post-ACS associates with bleeding risk (15), and acute myocardial infarction-induced coagulation activation alters peripheral vascular responses compared to stable CAD (21). Conventional coagulation markers (e.g., activated partial thromboplastin time, APTT), which correlates with clinical presentation in angiography patients (22) may complement GDF-15 in guiding CAD management. Despite parallels to other cardiovascular diseases, no studies have directly examined GDF-15 and coagulation in CAD.

This study is the first to investigate associations between GDF-15 and coagulation parameters (prothrombin time, (PT), APTT, fibrinogen, thrombin time (TT), D-dimer, and antithrombin III (AT3) in male CAD patients, including ACS and SA subgroups.

## Materials and Methods

### Study population and eligibility criteria

This retrospective cross-sectional study enrolled male patients who underwent elective coronary angiography for suspected CAD (including SA and ACS) at the Renmin Hospital of Wuhan University between January 2020 and December 2020. A control group of age-matched healthy males was also included. The study protocol was approved by the Medical Ethics Review Committee of Renmin Hospital, Wuhan University, and complied with the Declaration of Helsinki. Written informed consent was obtained from all participants.

Inclusion Criteria for CAD Patients: 1. Coronary angiography-confirmed stenosis≥50% occlusion in one major coronary artery or 30–50% occlusion with evidence of ischemia (resting or stress-induced). Exclusion Criteria: Severe comorbidities (e.g., active infection, pulmonary edema, chronic/acute kidney injury) or recent thrombolysis.

### Angiography Assessment

Two blinded radiologists evaluated stenosis severity using the Gensini score (23). Vessel disease was classified as 0 – 3 based on affected arteries (left anterior descending, left circumflex, right coronary). Left main trunk involvement was scored as 2-vessel disease.

### Definitions

In this study, SA is defined as a myocardial ischemia/hypoxia due to ≥50% coronary occlusion (24). ACS include ST-segment elevation myocardial infarction, non-ST-segment elevation myocardial infarction, and unstable angina per current guidelines (25).

### Laboratory measurements

Blood samples from acute myocardial infarction patients were collected immediately upon hospitalization due to the urgency of their condition. For other patients, venipuncture was performed in the morning after an overnight fast and prior to pharmacotherapy. Venous blood was drawn into plain tubes, centrifuged at 3500 rpm/min for 15 minutes at 25°C, and the separated plasma/serum was stored at −80°C until analysis. (leukocytes, neutrophils, monocytes, and lymphocytes)

Complete blood counts (white blood counts (WBC), neutrophils (Neu), monocytes (Mono), and lymphocytes (Lym)) were analyzed using a Sysmex XN-20 hematology analyzer (Kobe, Japan). Serum biochemical parameters (total cholesterol (TC), triglyceride (TG), high-density lipoprotein cholesterol (HDL-c), low-density lipoprotein cholesterol (LDL-c), and glucose) were quantified with an ADVIA 2400 biochemistry analyzer (Siemens, Germany). GDF-15 levels were measured via an Elecsys^®^ electrochemiluminescent immunoassay (Roche Diagnostics; research-use-only in China; detection range: 400–20,000 ng/L). Coagulation tests (APTT, PT, TT, fibrinogen) were performed on a CS-5100 analyzer (Sysmex, Japan) using the clotting method, while D-dimer (immunoturbidimetry) and AT3 (chromogenic substrate method) were assayed on the same platform.

All procedures strictly followed manufacturer-specified protocols and package insert guidelines.

### Statistical analysis

Continuous variables are described as median (interquartile range [IQR]), and categorical variables are summarized as frequencies (percentages) for baseline characteristics and laboratory parameters. After stratifying the entire CAD population into high, medium, and low GDF-15 groups based on tertiles (26), continuous variables were compared among the three groups using the nonparametric Kruskal-Wallis test. For significant omnibus tests (*P*<0.05), post hoc analyses were performed with the following approaches: (a) Holm-Sidak method for normally distributed data, or (b) Dunn’s test with Holm adjustment for non-normal distributions. All comparisons were conducted at a 5% overall significance level.

Categorical variables were analyzed using the Pearson chi-square test or Fisher’s exact test, as appropriate. For correlation analyses, the coefficient sign indicates the association direction (positive/negative), and its absolute value reflects the strength: 0.8–1.0 (very strong), 0.6–0.8 (strong), 0.4–0.6 (moderate), 0.2–0.4 (weak), and 0.0–0.2 (very weak). Multivariate linear regression assessed the relationship between plasma GDF-15 levels (log10-transformed to ensure normality of residuals) and coagulation parameters.

A two-sided *P* value <0.05 was considered statistically significant. Analyses were performed using IBM SPSS 23.0 (IBM Corp., Armonk, NY, USA) and GraphPad Prism 7.0 (GraphPad Software, La Jolla, CA, USA).

## Results

### Baseline characteristics

*In* this study, 892 subjects were enrolled, including 592 cases in the CAD group and 300 cases in the control group. The CAD group consisted of 161 SA cases and 431 ACS cases. As **Table 1** shown, no significant differences in age distribution and the incidence of hypertension and diabetes between CAD patients and healthy controls were determined (*P* > 0.05). However, the levels of inflammatory marks (WBC, Neu, and Mono), glucose, and lipids (TC, TG, HDL-c, and LDL-c) in the CAD group were higher compared to the control group (*P* <0.01). Additionally, the CAD group had significantly lower Lym counts. Notably, the levels of GDF-15 were significantly higher in the CAD group compared to the control group [700.00 (545.00-985.00) pg/mL vs. 1308.50 (890.00-1968.50) pg/mL, *P*<0.001].

**Table 1.**
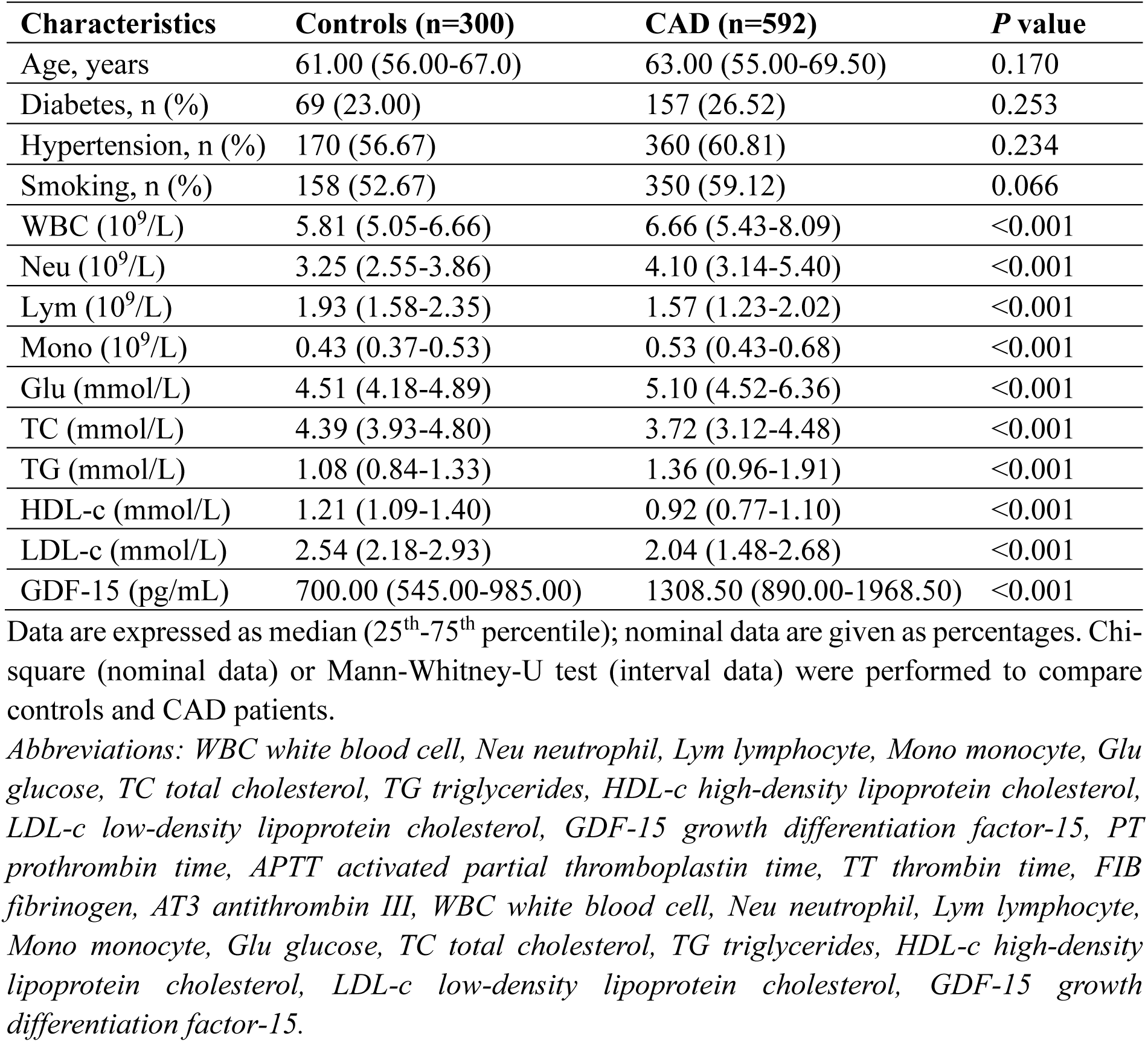
Baseline characteristics of the study population.

To further analyze the association between GDF-15 and various marks, we stratified CAD patients into tertiles based on GDF-15 level **(Table 2**), given 195, 196, 201 patients in low (<999.35 pg/mL), medium (999.35-1690.52 pg/mL), and high (≥1690.52 pg/mL) GDF-15 level subgroup, respectively. The higher tertile subgroups of GDF-15 showed higher proportions of patients with diabetes, hypertension, and higher levels of inflammatory marks (WBC, Neu, and Mono), and lipids (high TC, TG, and low HDL-c), as well as high Gensini Score. No significant differences in glucose and LDL-c concentrations were observed among the three subgroups (*P*>0.05). Regarding coagulation parameters, we observed differences among the three GDF-15 subgroups. Generally, PT and APTT were prolonged, fibrinogen and D-dimer levels were higher with increasing GDF-15 tertiles, while the activity of AT3 was significantly decreased.

**Table 2.**
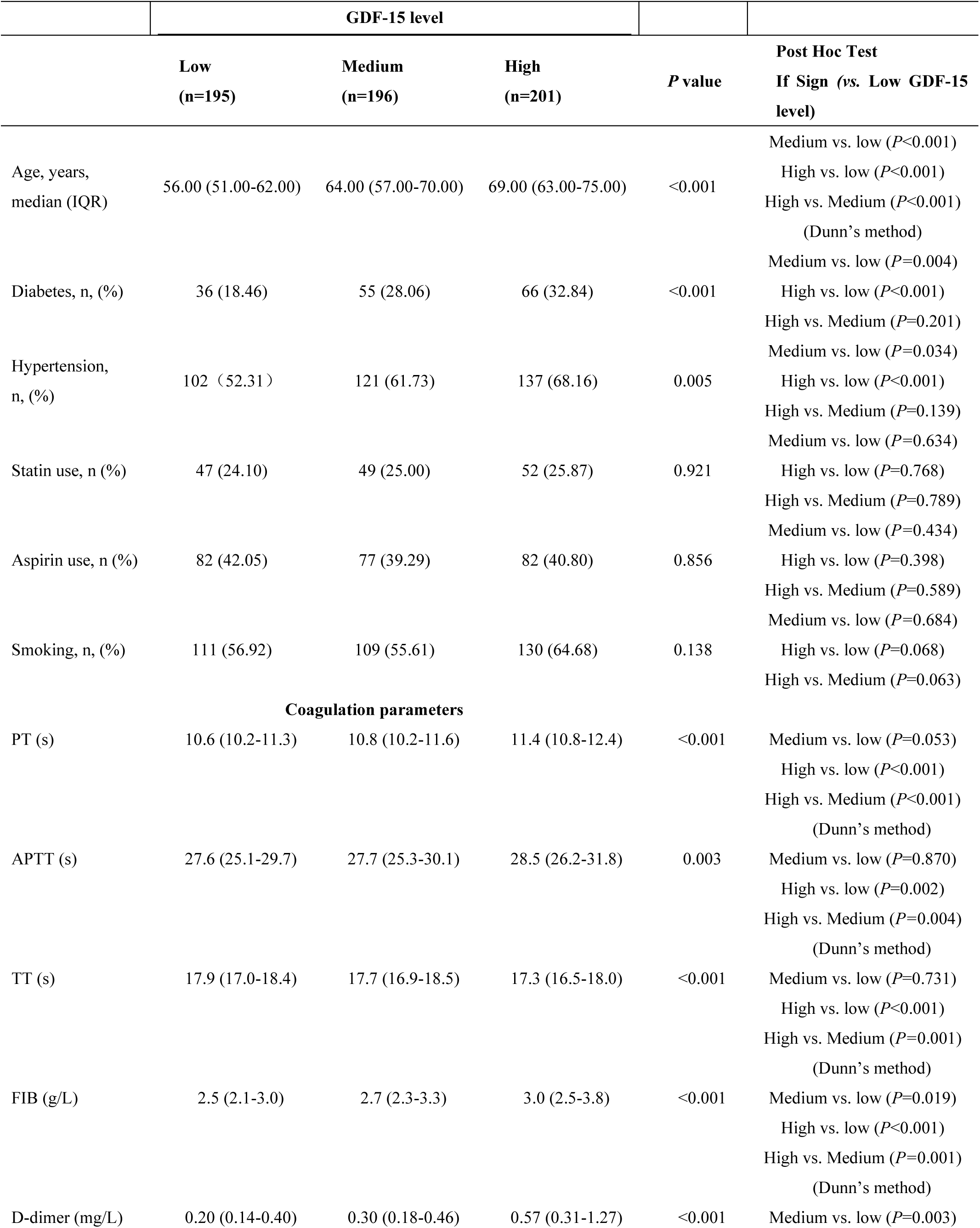
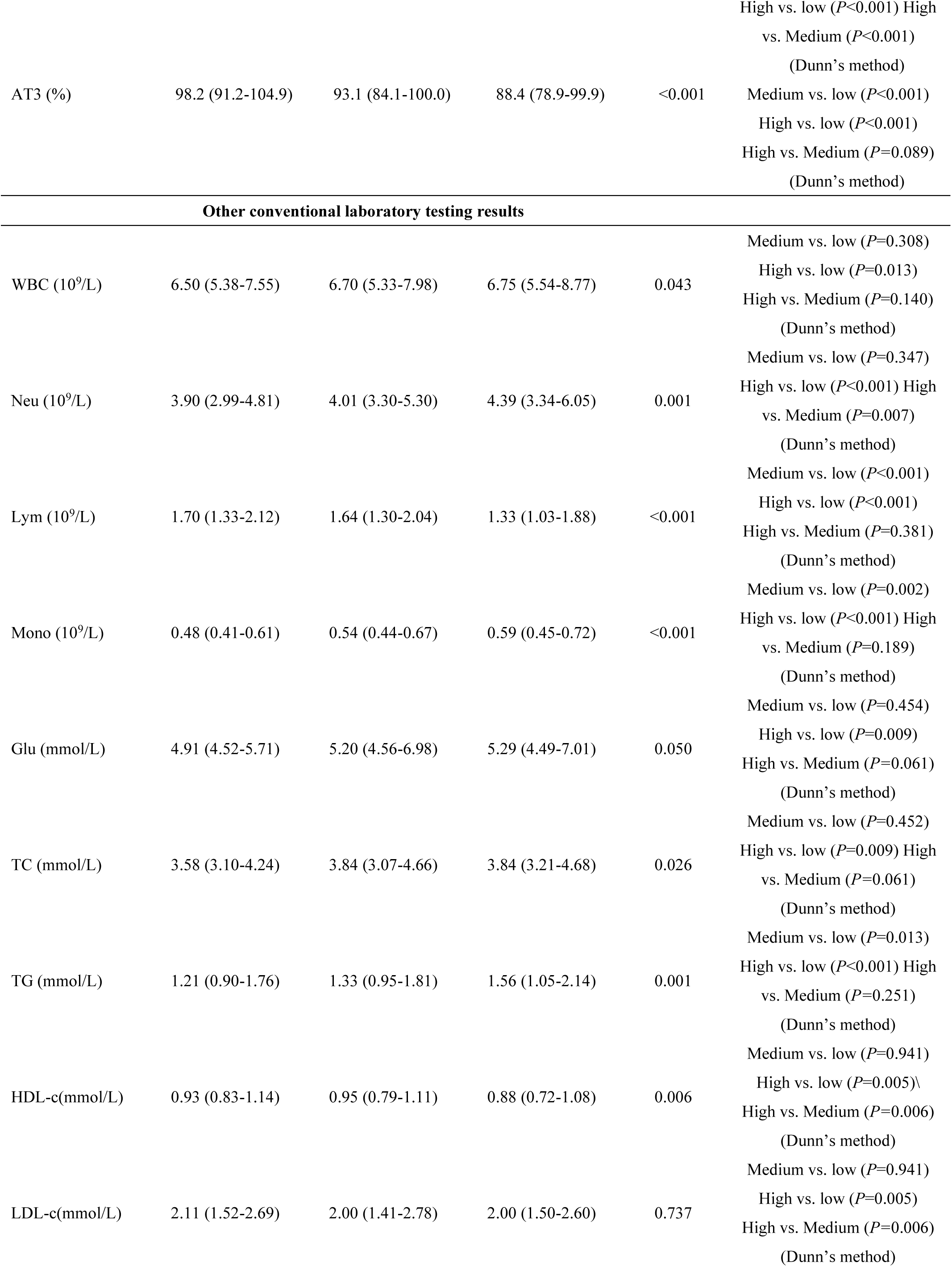
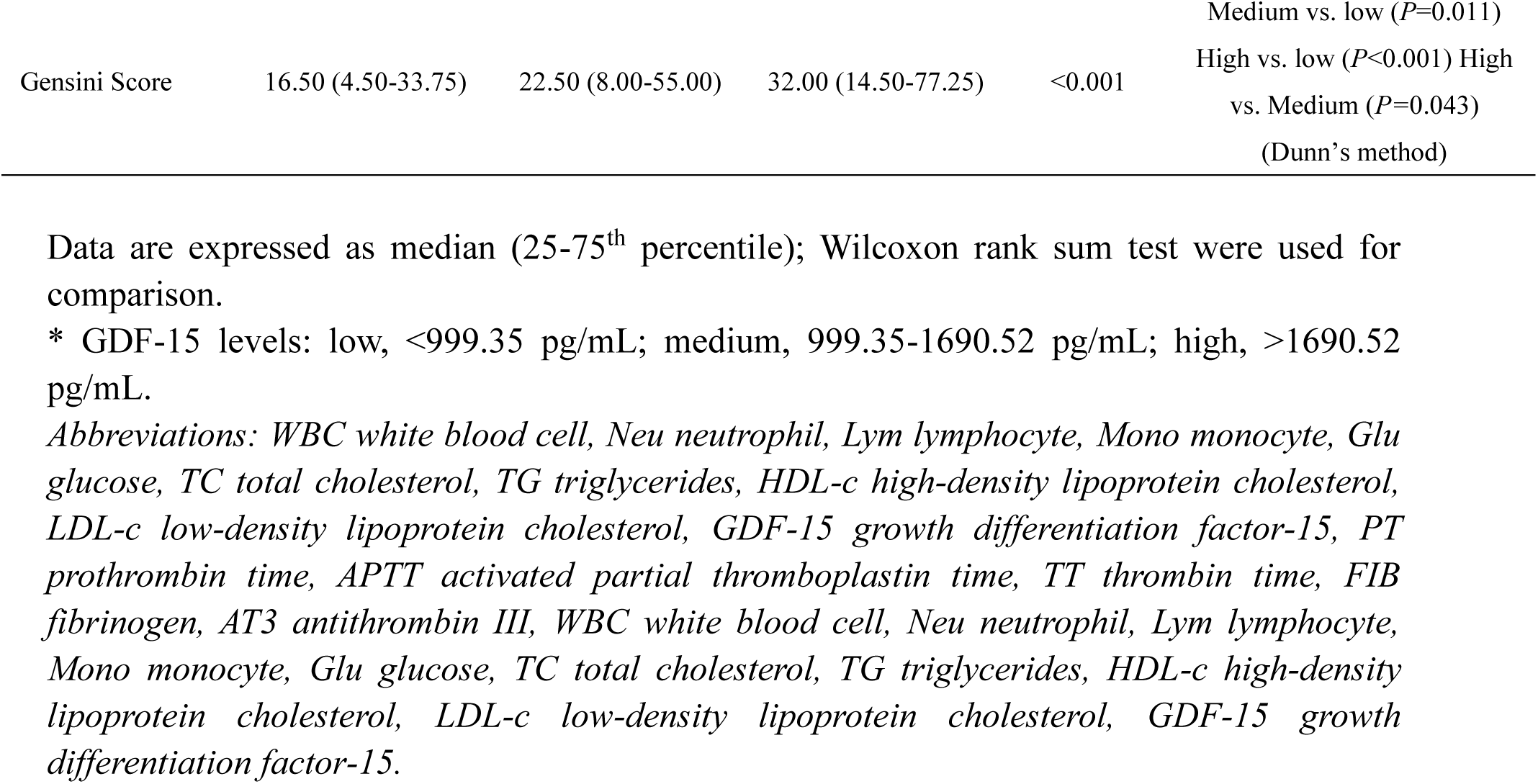
Baseline characteristics in relation to quartile groups of GDF-15 levels in CAD patients.

### Correlations of GDF-15 with coagulation parameters in CAD patients

**Table 3** demonstrates the correlations between GDF-15 levels and coagulation parameters. Weak and moderate correlations were observed between GDF-15 and APTT (*r*=0.159, *P*<0.001), fibrinogen (*r*=0.244, *P*<0.001), PT (*r*=0.337, *P*<0.001), and D-dimer (*r*=0.443, *P*<0.001). AT3 and TT levels were negatively and weakly correlated with GDF-15 (*r*= -0.335, *P*<0.001; *r*= -0.134, *P*=0.002).

**Table 3.** Correlations of GDF-15 level with coagulation parameters.

| Coagulation parameters | All CAD patients |  | SA (n=161) |  | ACS (n=431) |  |
| --- | --- | --- | --- | --- | --- | --- |
|  | *r* | *P* | *r* | *P* | *r* | *P* |
| PT | 0.337 | <0.001 | 0.052 | 0.521 | 0.353 | <0.001 |
| APTT | 0.159 | <0.001 | 0.042 | 0.609 | 0.227 | <0.001 |
| TT | -0.134 | 0.002 | -0.064 | 0.439 | -0.101 | 0.050 |
| FIB | 0.244 | <0.001 | 0.138 | 0.093 | 0.191 | <0.001 |
| D-dimer | 0.443 | <0.001 | 0.244 | 0.002 | 0.421 | <0.001 |
| AT3 | -0.335 | <0.001 | -0.281 | 0.001 | -0.257 | <0.001 |

*Abbreviations: GDF-15 growth differentiation factor-15, PT prothrombin time, APTT activated partial thromboplastin time, TT thrombin time, FIB fibrinogen, AT3 antithrombin III*

Subsequently, CAD patients were divided into SA and ACS subgroups for further sensitivity analysis. In the SA subgroup, significantly weak correlations were observed between GDF-15 and D-dimer (*r*=0.244, *P*=0.002) as well as AT3 (*r*= -0.281, *P*=0.001). In contrast, significant correlations were observed between GDF-15 and all coagulation parameters in patients with ACS, except TT . Specifically, weak correlations were observed between GDF-15 and TT (*r*= -0,101, *P*<0.001), fibrinogen (*r*=0.191, *P*<0.001), APTT (*r*=0.227, *P*<0.001), AT3 (*r*= -0.257, *P*<0.001), PT (*r*=0.353, *P*<0.001) and AT3 (*r*=-0.257, *P*<0.001). The correlation between GDF-15 and D-dimer is positive and moderate (*r*=0.421, *P*<0.001).

### Multivariate linear regression analyses on the associations of GDF-15 level with coagulation parameters

Multiple linear regression analyses were conducted to examine the associations between GDF-15 levels as the dependent variable and coagulation parameters as the independent variables in all participants and the SA and ACS subgroups. A negative multivariable coefficient indicated a negative association, and vice versa. As demonstrated in **Table 4**, GDF-15 showed a significant association with fibrinogen in all CAD patients and in the SA and ACS groups (all *P* < 0.05). Furthermore, GDF-15 was found to be significantly associated with AT3 in the ACS subgroup (*P*=0.039).

**Table 4.** Multivariate linear regression analyses on the associations of GDF-15 level with coagulation parameters.

| Independent variables | All CAD patients |  |  | SA |  |  | ACS |  |  |
| --- | --- | --- | --- | --- | --- | --- | --- | --- | --- |
| | *B* | $\beta$ | *P* | *B* | $\beta$ | *P* | *B* | $\beta$ | *P* |
| PT | 0.007 | 0.078 | 0.078 | -0.004 | -0.081 | 0.353 | 0.008 | 0.107 | 0.066 |
| APTT | 0.003 | 0.052 | 0.243 | 0.001 | 0.007 | 0.941 | 0.005 | 0.093 | 0.129 |
| TT | -0.003 | -0.069 | 0.074 | -0.001 | -0.119 | 0.137 | 0.001 | -0.005 | 0.925 |
| FIB | 0.053 | 0.208 | **<0.001** | 0.038 | 0.171 | **0.035** | 0.035 | 0.163 | **0.006** |
| D-dimer | 0.001 | 0.007 | 0.858 | -0.004 | -0.119 | 0.127 | 0.004 | 0.040 | 0.430 |
| AT3 | 0.001 | -0.069 | 0.083 | 0.001 | -0.142 | 0.080 | -0.002 | -0.120 | **0.039** |

\*Analyses were adjusted for age, hypertension, diabetes, and smoking status.
Abbreviations: *B*: Unstandardized coefficient; $\beta$ : Standardized coefficient, PT prothrombin time, APTT activated partial thromboplastin time, TT thrombin time, FIB fibrinogen, AT3 antithrombin III.

## Discussion

CAD ranks as the third leading cause of global mortality, characterized by poor prognosis. Its pathogenesis primarily involves excessive lipid deposition in vascular walls, which triggers immune dysfunction and aberrant release of inflammatory mediators (27). Although recent advancements in healthcare have contributed to a modest decline in both the incidence and mortality rates of CAD, it remains a predominant threat to global health (28). Importantly, CAD is not merely an age-related condition but rather a chronic inflammatory process. This process can precipitate acute clinical events through atherosclerotic plaque rupture or erosion, ultimately leading to arterial thrombosis. Given that thrombus formation is a life-threatening complication in CAD patients, early identification and intervention are critical for improving clinical outcomes. Consequently, investigating novel CAD biomarkers—including GDF-15 may provide valuable insights into the mechanisms underlying thrombus formation.

GDF-15, a cytokine typically expressed at low levels in various organs (including the liver, lungs, and kidneys), is upregulated in chronic diseases (29). A substantial body of research indicates that elevated plasma GDF-15 levels are associated with cardiovascular disease development, including CAD (30–32). The protein likely contributes to increased disease risk through multiple mechanisms, such as promoting CCR2-mediated macrophage chemotaxis toward atherosclerotic plaques and interleukin-6-dependent inflammatory responses (33). In this study, the CAD group exhibited significantly higher levels of inflammatory marks (WBC, Neu, and Mono), glucose, and lipids (TC, TG, HDL-c and LDL-c) compared to controls (*P* < 0.01), whereas lymphocyte (Lym) counts were lower. These differences in inflammatory markers may reflect GDF-15 upregulation in response to inflammatory or stress stimuli (34). Additionally, Additionally, elevated plasma GDF-15 correlated with adverse lipid profiles, consistent with prior studies linking GDF-15 to dyslipidemia (31, 35). Consistent with previous findings, our study confirmed higher GDF-15 in CAD patients than in healthy controls. Furthermore, when CAD patients were stratified by GDF-15 tertiles, high tertiles were associated with older age, diabetes, and hypertension—all established risk factors for CAD complications and mortality (36).

The pathogenesis of CAD involves a combination of extensive lipid deposition in the intima, endothelial dysfunction, exacerbated immune responses, proliferation of vascular smooth muscle cells, and extracellular matrix remodeling, ultimately leading to the formation of an atherosclerotic plaque (37). Coagulation and anticoagulation functions in patients with CAD are complex and influenced by multiple factors. Moreover, experimental and clinical studies have demonstrated the key role played by acute arterial thrombosis in the majority of myocardial infarctions and approximately 80% of strokes, collectively representing the leading cause of death in the developed countries (38). Rupture or damage of lipid-rich coronary plaques triggers subsequent thrombosis, especially atherothrombotic event, which is considered a crucial mechanism underlying the onset of ACS and ischemic sudden death (39). Advanced atherosclerotic plaques are characterized by the presence of a necrotic core predominantly composed of macrophages and vascular smooth muscle cells. The expansion of the necrotic core is also influenced by impaired clearance of necrotic cells, a process known as efferocytosis, leading to the accumulation of inflammatory material that further exacerbates atherosclerotic lesions, ultimately resulting in plaque rupture or erosion (40, 41).

In addition to its associations with inflammation and stress, elevated GDF-15 levels have been independently linked to an increased risk of stroke, systemic embolic events, and major bleeding (42–44). Notably, Siegbahn et al. found that higher GDF-15 concentrations significantly predicted major bleeding events in anticoagulated atrial fibrillation patients (45). Furthermore, Matusik et al. demonstrated that elevated GDF-15 independently predicts impaired fibrin clot lysability in AF patients, possibly due to its association with prothrombotic blood alterations (46). Together, these studies highlight GDF-15’s role in coagulation.

In our study, patients with higher serum GDF-15 exhibited prolonged PT, APTT, and TT, along with elevated fibrinogen and D-dimer levels and reduced AT3 activity. PT and APTT reflect exogenous and endogenous coagulation pathway dysfunction, respectively. Prolonged PT suggests deficiencies in vitamin K-dependent factors (II, VII, IX, X) or factor V. Prolonged APTT indicates impaired intrinsic pathway factors (VIII, IX, XI). D-dimer, a marker of fibrin degradation, aids in diagnosing thrombotic events (47). Fibrinogen, a key component of atherosclerotic plaques (48), contributes to thrombosis, inflammation, and blood viscosity. Giuseppe et al. reported that GDF-15 independently correlates with platelet function and fibrinogen levels in healthy adults (49). Similarly, our study confirmed a positive association between GDF-15 and fibrinogen in all CAD patients, including stable angina SA and ACS subgroups. Notably, in ACS patients, GDF-15 showed an inverse relationship with AT3—a critical serine protease inhibitor whose heparin-bound form mediates anticoagulation (50). Unlike SA, ACS involves acute coronary occlusion, leading to coagulation-anticoagulation imbalance, suggesting GDF-15 may offer additional insights into coagulation status in ACS

Several limitations of this study should be acknowledged. First, a significant limitation is the lack of analysis and data regarding the relationship between GDF-15 levels and clinical outcomes, including survival and major adverse cardiovascular events (MACE). Second, the study exclusively enrolled male CAD patients. While this approach was adopted to control for gender-related confounding factors, it should be noted that significant gender differences exist in CAD prevalence, with males being more susceptible to developing CAD. Third, as a case-control study with a relatively small sample size, the present investigation can only demonstrate an association between GDF-15 and coagulation parameters, rather than establish a causal relationship. Future studies with larger populations or longitudinal designs are warranted to further elucidate this relationship.

### Conclusion

In summary, this study demonstrated significant associations between elevated plasma GDF-15 levels and multiple coagulation parameters, particularly fibrinogen and AT3. These findings suggest that GDF-15 may serve as a compensatory marker for coagulation parameter instability. The results underscore the potential clinical utility of GDF-15 as a novel biomarker for assessing coagulation status in CAD patients, especially in the ACS subgroup

## Data Availability

All data produced in the present study are available upon reasonable request to the authors.

## Author contributions

Huan Liu and Yongnan Lyu performed most of the experiments, collected the samples and wrote the manuscript. Yan Li and Wen Dai designed this study and performed statistical analysis. All authors approved the submission of the final manuscript.

## Availability of data and materials

All data generated or analyzed during this study are included in this article. Further studies should be directed toward the corresponding authors.

## Consent for publication

Not applicable.

## Ethics approval and consent to participate

This study was conducted in strict accordance with the Declaration of Helsinki and approved by the ethics committee of Wuhan University (No.WDRY2020-K001). All participants signed an informed consent form before enrollment

## Acknowledgement

Medical editing assistance was provided by Dr. Xue Wu funded by Roche Diagnostics (Shanghai) Limited.

